# Deep-learning-Assisted Photoacoustic and Ultrasound Evaluation for Pre-transplant Human Liver Graft Quality and Transplant Suitability

**DOI:** 10.64898/2026.04.13.26350786

**Authors:** Qinghao Zhang, Kaustubh Pandit, Yan Cui, Feng Yan, Nanchao Wang, Jingting Li, Alan Yao, Luca Menozzi, Kar-Ming Fung, Zhongxin Yu, Paige Parrack, Walid Ali, Ronghao Liu, Chen Wang, Junyuan Liu, Clint A. Hostetler, Ashley N. Milam, Bradon Nave, Ronald A. Squires, Narendra R. Battula, Chongle Pan, Paulo N. Martins, Junjie Yao, Tri Vu, Qinggong Tang

## Abstract

End-stage liver disease (ESLD) is one of the leading causes of death worldwide. Currently, the only curative option for patients with ESLD is liver transplantation. However, the demand for donor livers far exceeds the available supply, partly because many potentially viable livers are discarded following biopsy evaluation. While biopsy is the gold standard for assessing liver histological features related to graft quality and transplant suitability, it often leads to high discard rates due to its susceptibility to sampling errors and limited spatial coverage. Besides, biopsy is invasive, time-consuming, and unavailable in clinical facilities with limited resources. Here, we present an AI-assisted photoacoustic/ultrasound (PA/US) imaging framework for quantitative assessment of human donor liver graft quality and transplant suitablity at the whole-organ scale. With multimodal volumetric PA/US images as the input, our deep-learning (DL) model accurately predicted the risk level of fibrosis and steatosis, which indicate the graft quality and transplant suitability, when comparing with true pathological scores. DL also identified the imaging modes (PAI wavelength and B-mode USI) that correlated the most with prediction accuracy, without relying on ill-posed spectral unmixing. Our method was evaluated in six discarded human donor livers comprising sixty spatially matched regions of interest. Our study will pave the way for a new standard of care in organ graft quality and transplant suitability that is fast, noninvasive, and spatially thorough to prevent unnecessary organ discards in liver transplantation.

## Introduction

The human liver is responsible for numerous metabolic detoxification, and synthetic processes essential to sustaining life. However, liver diseases accounted for over two million deaths globally in 2021, making them the 11th most common cause of mortality^1^. End-stage liver diseases (ESLDs), including cirrhosis, acute liver failure, and hepatocellular carcinoma (HCC), are the most common frequent contributors^2–5^. Liver transplantation remains a life-saving procedure for patients with ESLDs, with approximately 35,000 transplants performed worldwide in 2021^2–5^. In the US alone, over 9,400 patients are currently on the waitlist for liver transplantation, and around 2,000 patients die each year while waiting^6, 7^. The demand for liver organs in the U.S. continues to outgrow the supply significantly with an 18% annual increase in procedures over the past five years^3^. Meanwhile mortality rate on the waitlist is approaching 25%^8^. Beyond the clinical toll, patients on the waitlist can face medical expenses of approximately $23,000 per month^9^. Although there have been advances in regenerative medicine and tissue engineering, living and deceased donor liver transplantation remains the primary treatment option for ESLDs^10, 11^. Globally, only 10% of patients in need of a transplant receive one, highlighting the urgency of the organ shortage crisis^1^.

A significant contributor to the shortage is underutilization of donor organs. Currently, in the U.S., 76% of potential donor livers are discarded, often due to concerns about quality^12^. The gold standard for assessing organ quality is biopsy, which examines fibrosis (excess extracellular matrix protein) and steatosis (excessive lipid accumulation)^8, 13^. These are critical predictors of post-transplant outcomes, graft function, and overall patient mortality^14, 15^. Generally, an organ is deemed acceptable if it shows less than 30% macro vesicular steatosis and only mild fibrosis (Grade 0 or 1)^13^. However, biopsy is invasive and prone to sampling error because of tissue heterogeneity, as well as intra- and inter-observer variability^16–18^. Consequently, up to 70% of discarded livers may have had acceptable macro steatosis levels (<30%)^8^. Therefore, there is a critical need for an improved gold standard-method to optimize donor organ usage.

Noninvasive imaging modalities, such as ultrasound, CT, and MRI, are used in both living and deceased donors to assess organ graft quality and transplant suitablity^19, 20^. Ultrasound provides rapid evaluation of liver stiffness for fibrosis^21, 22^. However, it lacks direct quantitative information about collagen and lipid content, which are central to assess fibrosis and steatosis^23, 24^. CT and MRI also cannot be easily deployed in the operating rooms (OR) for quick evaluation before transplantation. Hyperspectral imaging (HSI) can provide valuable functional data (e.g., steatosis level, lactate concentration) during normothermic machine perfusion (NMP)^25–27^. However, it remains limited by shallow optical penetration depth^26^. These shortcomings underscore the need for a portable imaging tool that delivers deep-tissue, spatially comprehensive molecular assessments of the entire liver.

Photoacoustic tomography (PAT) is a promising solution to fulfill these requirements. By combining short-pulse laser excitation with ultrasound detection, PAT harnesses the dependence on optical absorption to make it intrinsically sensitive to highly absorbing endogenous molecules^28–30^. Without contrast and labelling, PAT can readily image a variety of molecules found within tissue, including both collagen and lipid – the hallmark of fibrosis and steatosis, respectively^31, 32^. With acoustic detection, PAT noninvasively provides deep-tissue whole-organ scanning in minutes without being subjected to spatial heterogeneity and sampling error. With hardware similarity to commercial ultrasound scanners, PAT system can be seamlessly integrated into current clinical ultrasound workflow, providing mobile usage in the OR. Hence, PAT presents a promising and clinical-friendly solution for sensitive deep- and whole-organ functional imaging of molecular contents for liver transplant.

Despite its potential, clinical translation of PAT remains constrained by the limitations of conventional spectral unmixing methods^33–36^. While linear spectral unmixing is computationally efficient, its quantitative fidelity in biological tissues deteriorates due to the wavelength-dependent tissue attenuation^34, 37^. It causes local optical fluence to become non-linearly coupled to chromophore absorption, making the inversion of molecules’ concentrations intrinsically ill-posed, especially for collagen and lipid in our case^34, 37–40^. Nonlinear unmixing methods^41–43^, despite accounting for light fluence, are still vulnerable to spectral shifts by pathological conditions, such as cirrhosis or steatosis. Disease-induced microenvironmental perturbations, such as local pH fluctuations, temperature fluctuations, redox alterations generating dyshemoglobins and binding-state variations^37, 44–47^, can subtly alter chromophores’ electronic states and shift their absorption spectra. Combining with unmodeled^46^ endogenous absorbers, these effects make it challenging for existing unmixing algorithms to disentangle molecules reliable for organ assessment ^34, 37, 47^.

Meanwhile, ultrasound imaging (USI), especially B-mode, has been widely adopted for liver fibrosis^48–50^. Using echogenicity and speckle heterogeneity, B-mode USI can provide structural information related to fibrosis, but lacks direct sensitivity to underlying tissue composition.. However, there is a lack of methods to combine both PAT and USI effectively to provide quantitative assessment. Although PAT and USI are often acquired together, they are usually analyzed separately^51–53^. USI is mainly used to evaluate structural features, while PAT provides molecular information. However, these two modalities are rarely combined into a unified quantitative framework for evaluating liver pathology. Even when multimodal approaches are explored, they are typically implemented through feature extraction rather than direct data integration. Feature-based approaches may fail to capture critical information required for accurate prediction. This process reduces high-dimensional spatial and spectral data into a limited set of statistical values and may overlook spatial patterns and interactions within the intrinsic data^54, 55^. Thus, there is a need for an effective method that takes multi-channel PAT and USI and outputs assessment outcomes, which fully utilizes the spatial and spectral information of the multi-modality data.

Meanwhile, the use of AI and deep learning (DL) has significantly advanced the field of medical imaging. First, it aids physicians in making quick and accurate diagnostic decisions in medical imaging, which is particularly important for organ quality and transplant suitability. AI tools are increasingly used in clinical imaging to support screening and diagnosis and have been shown to reduce workload and improve efficiency^55, 56^. Similar strategies have been explored in organ assessment prior to transplantation^57^. Second, DL helps resolve ill-posed inverse problems in imaging modalities that depend heavily on physical models, especially PAT and USI. Instead of relying solely on explicit physical inversion, DL learns the mapping between input measurements and target outputs directly from data. This data-driven approach has been applied to address ill-posed reconstruction and unmixing problems in imaging^58–61^. DL also identifies underlying features and characteristics in images that lead to better prediction, especially in USI. In contrast to handcrafted feature extraction, DL performs end-to-end learning and automatically derives task-specific features from raw images^62–64^. Thus, there is significant potential of using AI/DL for PAT/USI evaluation of organ quality and transplant suitability before transplantation.

In this study, we developed a dual-modality 3D PAT/USI framework with deep learning to assess discarded human donor livers using spatially matched histopathology as ground truth. The model directly predicts clinically relevant steatosis and fibrosis risk from multimodal imaging data. Multispectral PAT volumes (680–970 nm) and co-registered B-mode ultrasound were directly input to DL model to decide on organ quality and transplant suitability before transplantation. Our method bypasses spectral unmixing, enabling end-to-end learning from raw spatial–spectral information and avoiding inversion-related assumptions. DL models were evaluated using leave-one-donor-out cross-validation to enforce donor-level generalization. In 60 matched ROIs from six donor livers, multispectral PAT improved steatosis discrimination, whereas fibrosis discrimination relied primarily on B-mode ultrasound. Whole-organ prediction maps further demonstrated that the model could preserve regional patterns in overall intermediate-risk livers with heterogeneous pathology, supporting organ-scale evaluation beyond the sampling limits of biopsy. This framework provided a clinically feasible strategy to reduce biopsy-dependent uncertainty in pre-transplant liver assessment.

## Methods

### Donor organ handling and experimental setup for ex vivo organ imaging

All human liver specimens were obtained from LifeShare of Oklahoma, a non-profit Organ Procurement Organization (OPO) dedicated to transplantation purposes (**Fig. 1b-iii**). Following retrieval from LifeShare of Oklahoma OPO, the organs were then prepared and imaged using PAI system within 48 hours (**Fig. 1b-ii, iii, c**). The study is approved by IRB protocol (12462). A total of ten donor livers were imaged, of which six had spatially matched histological labels and were included in the supervised evaluation. Each liver underwent standardized preparation including thorough cleaning with deionized water to remove surface debris. A custom-designed water chamber filled with deionized water was placed over the organ for two purposes. First, it served as the acoustic coupling medium to ensure efficient transmission of acoustic signals between the liver surface and the transducer. Second, it ensures flat and long area of liver scanning.

**Figure 1.**
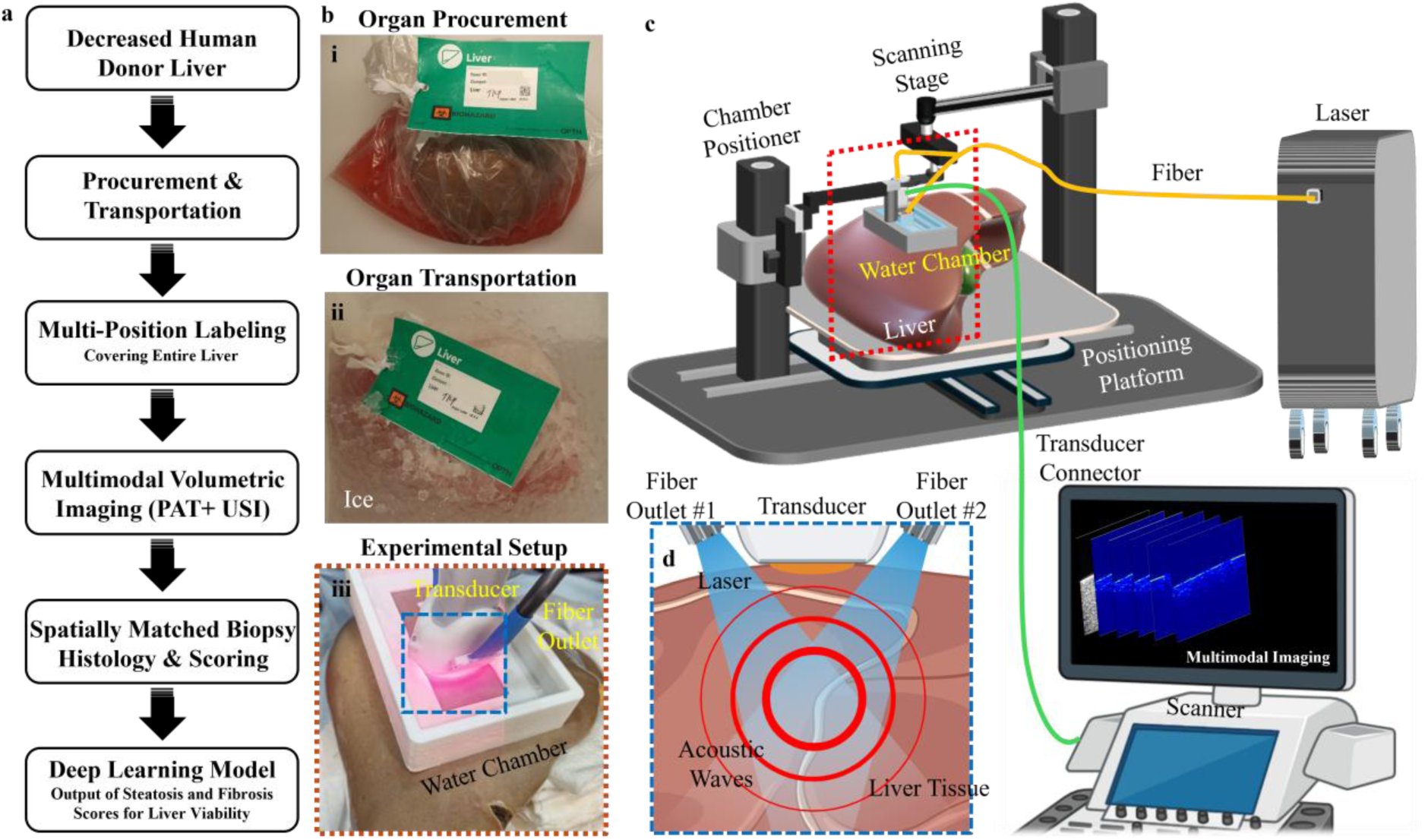
PAT and USI evaluation of liver transplant. (a) Workflow of the liver quality and transplant suitability assessment using PAT and USI. (b) Representative images of the organ at selected steps, including (i) after procurement, (ii) after transportation to our research facility, and (iii) during PAT/USI scan. (c) Full schematics of the experimental setup in (b-iii). (d) Zoomed-in schematics of the liver PAT/USI.

### Organ-scale volumetric PAT/US imaging workflow

The overall experimental workflow was illustrated in **Fig. 1a**. Discarded human donor livers were first procured and transported to imaging room. To capture organ-scale heterogeneity, multiple spatially distributed regions across the liver cortical surface were marked prior to imaging. Multimodal volumetric imaging (PAT and B-mode ultrasound) was then performed at each marked location, followed by spatially matched biopsy sampling for histological scoring. These region-level imaging volumes and pathology scores were subsequently used to train and evaluate a deep learning model for predicting steatosis and fibrosis risk.

Volumetric PAT imaging was performed using a dual-modality PAT/ultrasound system (Vevo F2 LAZR-X, Fujifilm VisualSonics) (**Fig. 1c**). The system contains two main parts. First, for PAT signal generation, it uses a short-pulsed laser (10-ns pulse width, 20-Hz repetition rate) with tunable wavelength from 680-970 thanks to the optical parametric oscillator. Five excitation wavelengths (680, 730, 860, 926, and 970nm) were selected to span lipid, collagen, and hemoglobin-dominant absorption regimes, enabling multispectral characterization of hepatic tissue without relying on linear spectral inversion. The laser’s peak energy is 50 mJ at 750 nm over a large illumination area (∼3 cm^2^), corresponding to an energy density of 16.7 mJ/cm^2^. Thus, it is well within the laser safety limit of 25 mJ/cm^2^ ^65^. The laser was delivered to the sample by a fiber bundle with a dark-field illumination (**Fig. 1c, d**). Second, for ultrasonic signal generation and acquisition, we use a linear-array transducer (UHF29x, 20-MHz center frequency, 23-mm lateral width). The transducer was mounted on a linear translational stage. The scanning protocol covered approximately 75% of the liver cortical surface. The sequential scans acquired at 0.15 mm step sizes across the marked regions, which ensured sufficient lateral sampling density for volumetric reconstruction and avoided redundant oversampling. At each scan, PAT images from five wavelengths and ultrasound B-mode were recorded. Following ultrasonic detection, raw RF data was beamformed using delay-and-sum.

### Histology & pathology

Histological analysis was performed on spatially matched liver tissue samples to provide gold-standard validation of imaging findings. Tissue samples were excised from marked imaging locations using a standardized protocol to ensure spatial correspondence between imaging and histology. Each sample was fixed in 10% neutral-buffered formalin, followed by routine tissue processing, paraffin embedding, and sectioning. Multiple staining techniques were employed to comprehensively assess liver tissue quality and transplant suitability markers. Hematoxylin and eosin (H&E) staining provided overall morphological assessment, while Masson’s trichrome staining specifically highlighted collagen deposition for fibrosis evaluation^66, 67^. All stained sections were digitized using a high-resolution slide scanner to facilitate quantitative image analysis and comparison with imaging data. The histological specimens were evaluated independently by a board-certified pathologist who was blinded to imaging results and specimens were scored for fibrosis and steatosis. For deep learning classification, pathology scores were binarized into low-and high-risk groups. For steatosis, Grades 0–1 (≤33%) were defined as low risk and Grades 2–3 (>33%) as high risk^68, 69^. For fibrosis, Grades 0–1 were defined as low risk and Grades 2–4 as high risk^70^.

### Volumetric Deep Learning Analysis

The overall deep learning framework for organ-scale risk prediction was illustrated in **Fig. 2a**. Multispectral PAT volumes acquired at five wavelengths (680, 730, 860, 926, and 970 nm) were co-registered with B-mode ultrasound volumes and concatenated to form six-channel three-dimensional inputs (**Fig. 2b**). Linear spectral unmixing was not performed; instead, the original multispectral signals were retained to avoid additional assumptions associated with inversion-based quantification.

**Figure 2.**
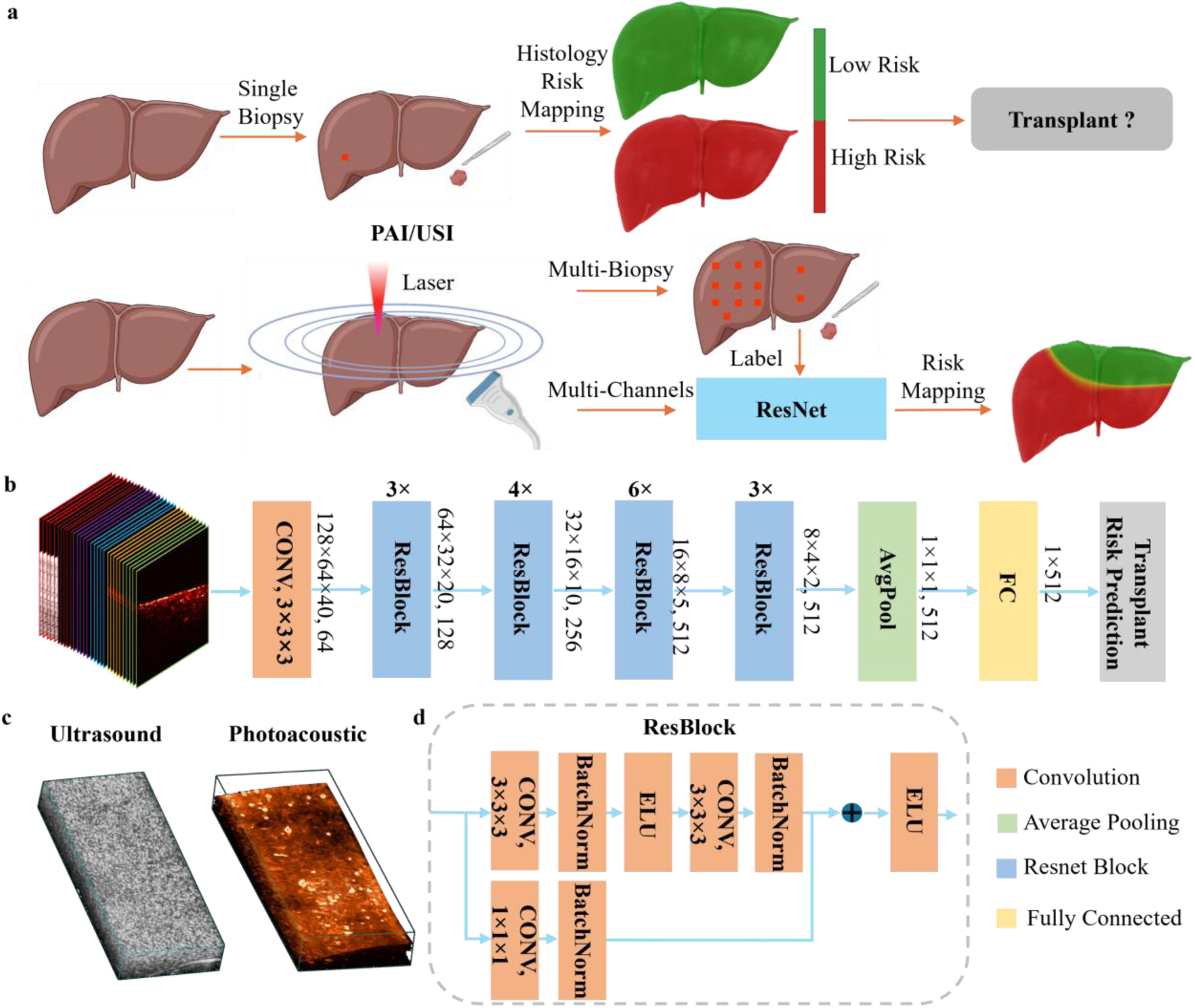
Organ-scale risk prediction using multispectral PAT/USI and volumetric deep learning. (a) Schematic workflow. Multispectral PAT/USI is performed at multiple positions across the donor liver. Spatially matched biopsies provide regional labels for model training. The trained network generates organ-level risk maps. (b) Architecture of the three-dimensional residual network used for classification. Six-channel volumetric inputs (five PAT wavelengths and B-mode ultrasound) are processed through residual blocks followed by global average pooling and a fully connected layer. (c) Representative volumetric ultrasound and photoacoustic images. (d) schematic of a residual block.

Each ROI volume was divided into eight non-overlapping three-dimensional patches (2×2×2 grid). Each patch was resized to 128 × 80 × 256 voxels (approximately 1.15 × 1.6 × 0.75 cm) to standardize the spatial resolution across donor organs. A three-dimensional residual network was used for binary classification^62, 71, 72^. The architecture comprised four residual stages with progressively increasing channel dimensions (64–512), followed by global average pooling and a fully connected output layer.

Because the number of labeled donor livers was limited, self-supervised pretraining was applied to improve model initialization. Pretraining used volumetric PAT/US data from all ten donor livers (including four livers without pathologist labels) and supervised leave-one-donor-out training, and evaluation was performed only on the six livers with matched pathologist histology. Volumetric PAT/USI data from ten donor livers were used in a rotation-prediction task, and the pretrained encoder weights were subsequently fine-tuned using pathologist-labeled liver data.

Model performance was evaluated using leave-one-donor-out cross-validation across six labeled livers to assess generalization across organs. In each fold, five donors were used for training and one donor for testing. All normalization, augmentation, and class balancing procedures were applied to training data only. Performance was primarily assessed using the area under the receiver operating characteristic curve (AUC).

Sensitivity, specificity, and balanced accuracy were calculated at the optimal threshold determined by Youden’s index. Statistical comparisons of model performance were performed using unpaired two-sided t-tests on fold-level AUC values obtained from leave-one-donor-out cross-validation. A p-value < 0.05 was considered statistically significant.

## Results

### Multimodal Performance for Steatosis and Fibrosis

Multispectral PAT improved discrimination of hepatic steatosis compared with single-modality ultrasound. As summarized in **Fig. 3a**, the combination of all wavelengths with B-mode achieved the highest AUC for steatosis classification (0.878). This performance was significantly higher than that of B-mode ultrasound alone (AUC = 0.786, p = 0.016, two-tailed t-test). Reduced multimodal configurations also showed strong performance. In particular, the combination of 860 nm + 926 nm + B-mode achieved an AUC of 0.871, while 860 nm + B-mode achieved an AUC of 0.863. Both configurations showed no statistically significant difference compared with the full multispectral configuration (p > 0.05). These results indicate that comparable discrimination performance can be achieved with fewer wavelengths.

**Figure 3.**
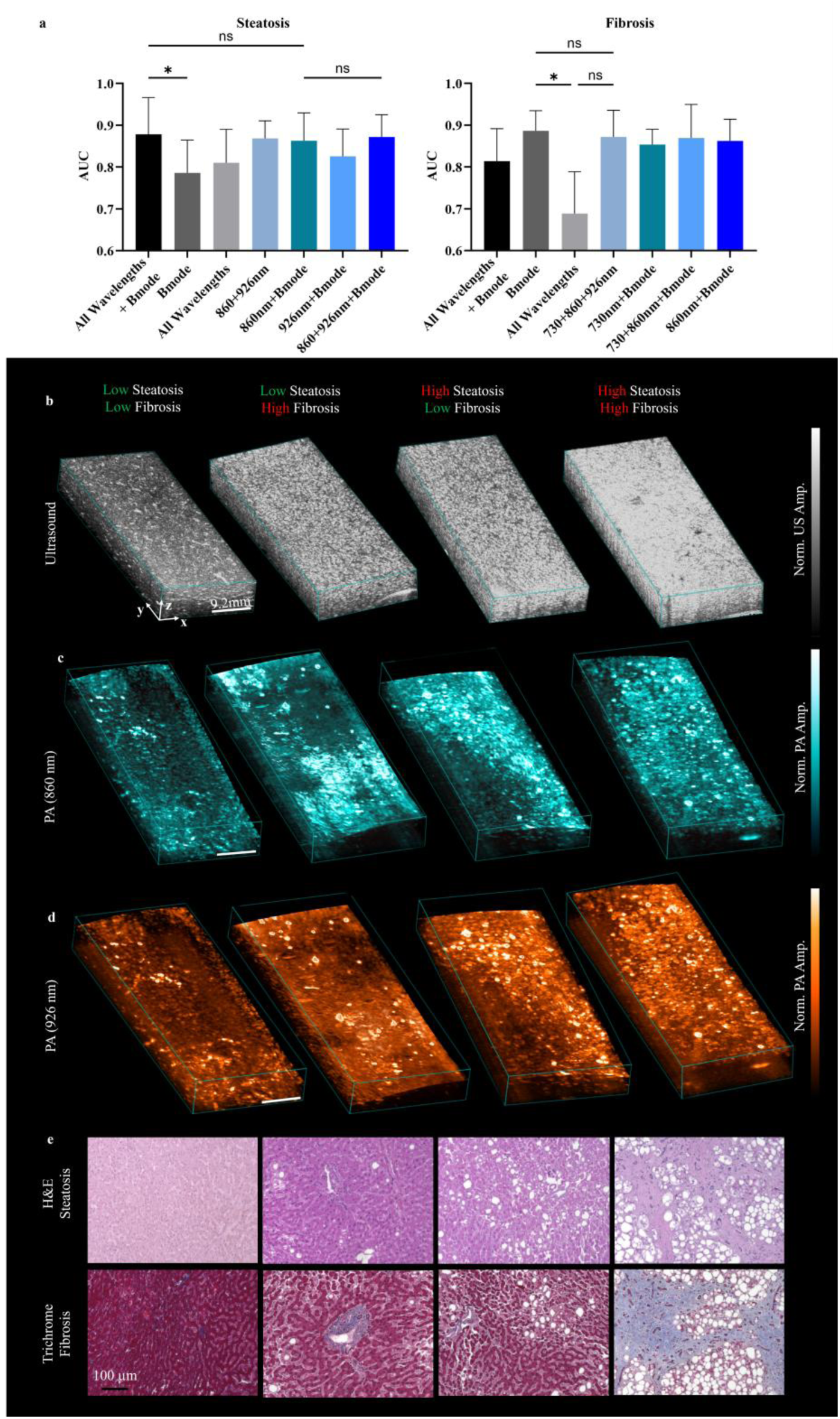
Classification performance and representative imaging findings for steatosis and fibrosis. (a) Comparison of AUC values across different wavelength and modality combinations for steatosis and fibrosis classification. Statistical significance is indicated. (b–d) Representative volumetric ultrasound and PA images (860 nm and 926 nm) from low- and high-risk samples. (e) Corresponding histological sections (H&E and Masson’s trichrome) confirming steatosis and fibrosis severity.

Across individual PAT wavelengths, AUC values ranged from 0.766 to 0.838. Among the single-wavelength configurations, 926 nm produced the highest performance (0.838), whereas shorter wavelengths such as 680 nm (0.766) showed comparatively lower discrimination ability and had significant difference with 926 nm (p = 0.026). PAT-only wavelength combinations demonstrated intermediate performance. For example, the three-wavelength combination of 730 nm + 860 nm + 926 nm achieved an AUC of 0.832, while the dual-wavelength combination of 860 nm + 926 nm achieved an AUC of 0.868. When B-mode ultrasound was combined with subsets of PAT wavelengths, AUC values generally ranged between 0.795 and 0.871 depending on the wavelength combination. Detailed AUC values for individual wavelengths and all wavelength combinations are summarized in **Supplementary Table 1**.

For fibrosis classification, B-mode ultrasound alone achieved the highest AUC (0.886). In comparison, individual PAT wavelengths yielded lower AUC values ranging from 0.686 to 0.797, with 860 nm showing the strongest PAT-only performance. PAT-only wavelength combinations demonstrated moderate discrimination ability. For example, the three-wavelength combination of 730 nm + 860 nm + 926 nm achieved an AUC of 0.872, which was not significantly different from B-mode ultrasound (p > 0.05). When B-mode ultrasound was combined with subsets of PAT wavelengths, the resulting AUC values ranged from 0.847 to 0.869 across different configurations but did not consistently outperform ultrasound alone.

Considering that fewer wavelengths reduce acquisition time, the 860 nm + B-mode configuration provides a practical balance between diagnostic performance and imaging efficiency for steatosis assessment. For fibrosis, B-mode ultrasound alone achieved the highest discrimination performance within the evaluated configurations.

Representative examples are shown in **Fig. 3b–d**. High-steatosis samples demonstrated globally elevated PAT intensity at both 860 nm and 926 nm compared with low-risk regions. In contrast, fibrosis-related differences were less prominent in the multispectral PAT images within the 680–970 nm range.

Corresponding histological sections (**Fig. 3e**) confirmed marked macrovesicular fat accumulation on H&E staining and increased collagen deposition on Masson’s trichrome staining, validating the imaging-based classification for both steatosis and fibrosis.

### Cross-Validation Performance and Whole-Organ Prediction Maps

Steatosis discrimination remained consistent across leave-one-donor-out folds (**Fig. 4a**), with multispectral PAT combined with B-mode ultrasound achieving stable separation between low- and high-risk regions and limited inter-fold variability. This indicates that the model maintained robust performance across different donor organs despite inter-donor heterogeneity in tissue composition and pathology. For fibrosis, B-mode ultrasound alone demonstrated the highest and most consistent performance across folds, indicating that fibrosis prediction in this dataset was primarily driven by structural acoustic features, with limited contribution from multispectral PAT within the examined spectral range.

**Figure 4.**
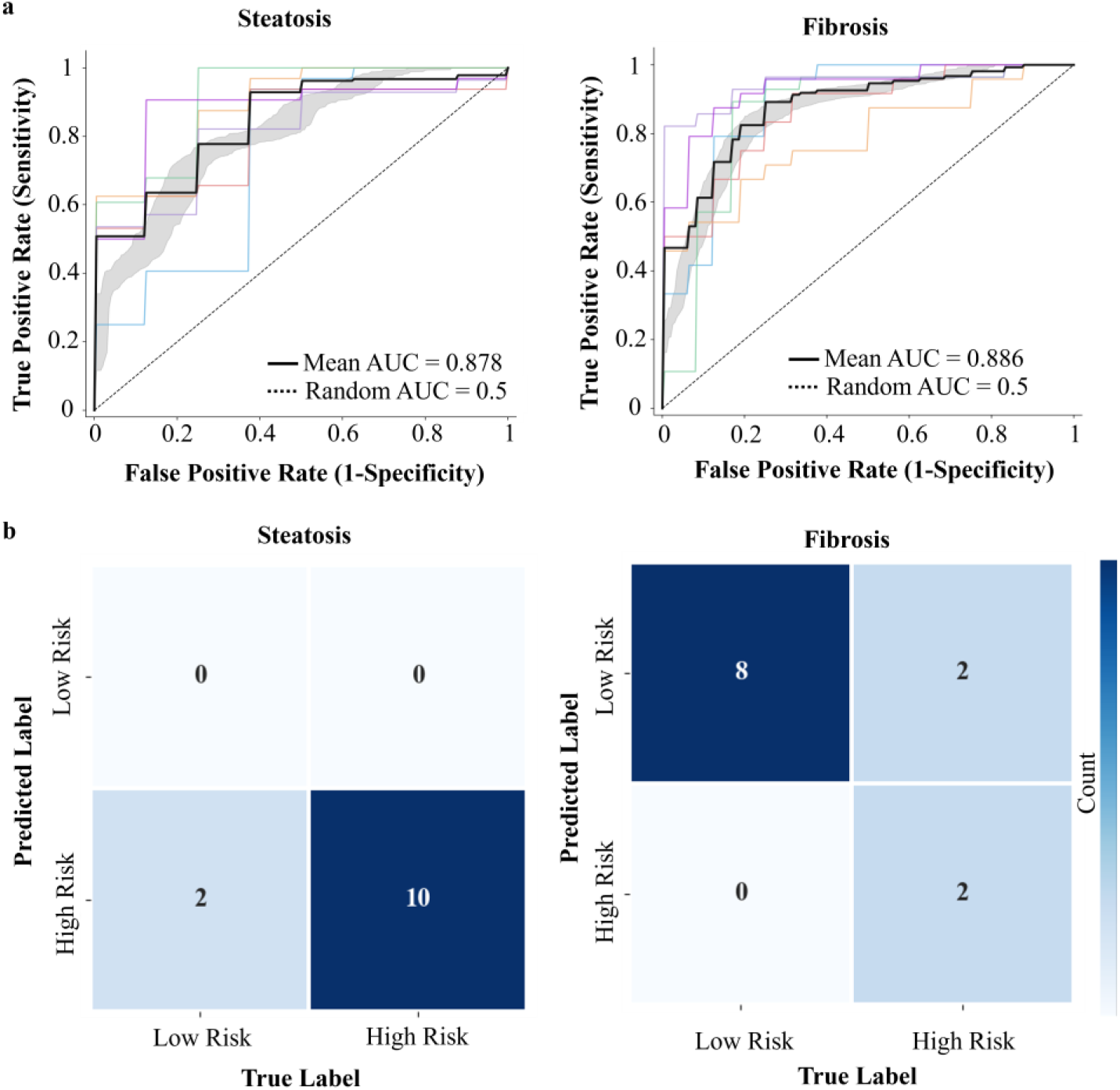
Cross-validation performance for liver steatosis and fibrosis. (a) Representative ROC curves across leave-one-donor-out folds for steatosis (all wavelengths + B-mode) and fibrosis (B-mode), with each curve corresponding to one of the six donor livers. (b) Corresponding confusion matrices for the selected modality combinations. Steatosis (All wavelengths + B-mode) and fibrosis (B-mode).

Confusion matrices (**Fig. 4b**) revealed distinct error patterns. For steatosis, most high-risk regions were correctly identified, with no false negatives and 2 observed false positives, indicating high sensitivity with a slight reduction in specificity. This asymmetric error distribution suggests a conservative prediction tendency toward high-risk classification, likely related to the imbalanced distribution of steatosis in the dataset, where high-risk regions were more prevalent. For fibrosis, high-risk regions were consistently detected, with misclassifications arising mainly from overestimation in low-risk regions, indicating reduced specificity. Compared with steatosis, the more balanced label distribution in fibrosis resulted in less pronounced prediction bias, although structural feature overlaps between low- and high-risk regions may still contribute to false positives.

Should add a subsection title here,

Whole-organ prediction maps (**Fig. 5**) demonstrated spatially coherent risk patterns across overall low, intermediate, and high-risk livers. For steatosis, spatial agreement was high in high-risk and low-risk cases; however, intermediate-risk livers with widespread fat involvement showed a tendency toward overprediction, resulting in partial loss of spatial heterogeneity (**Fig.5a-c**). This behavior is consistent with the confusion matrix trends and reflects the imbalanced composition of the dataset, where the dominance of steatotic regions may bias the model toward globally elevated risk predictions, particularly in heterogeneous organs. Fibrosis maps (**Fig.5d, e**) closely matched the ground-truth distributions, capturing both inter-organ differences in overall severity and intra-organ spatial variability, with regional heterogeneity particularly well preserved in intermediate-risk livers.

**Figure 5.**
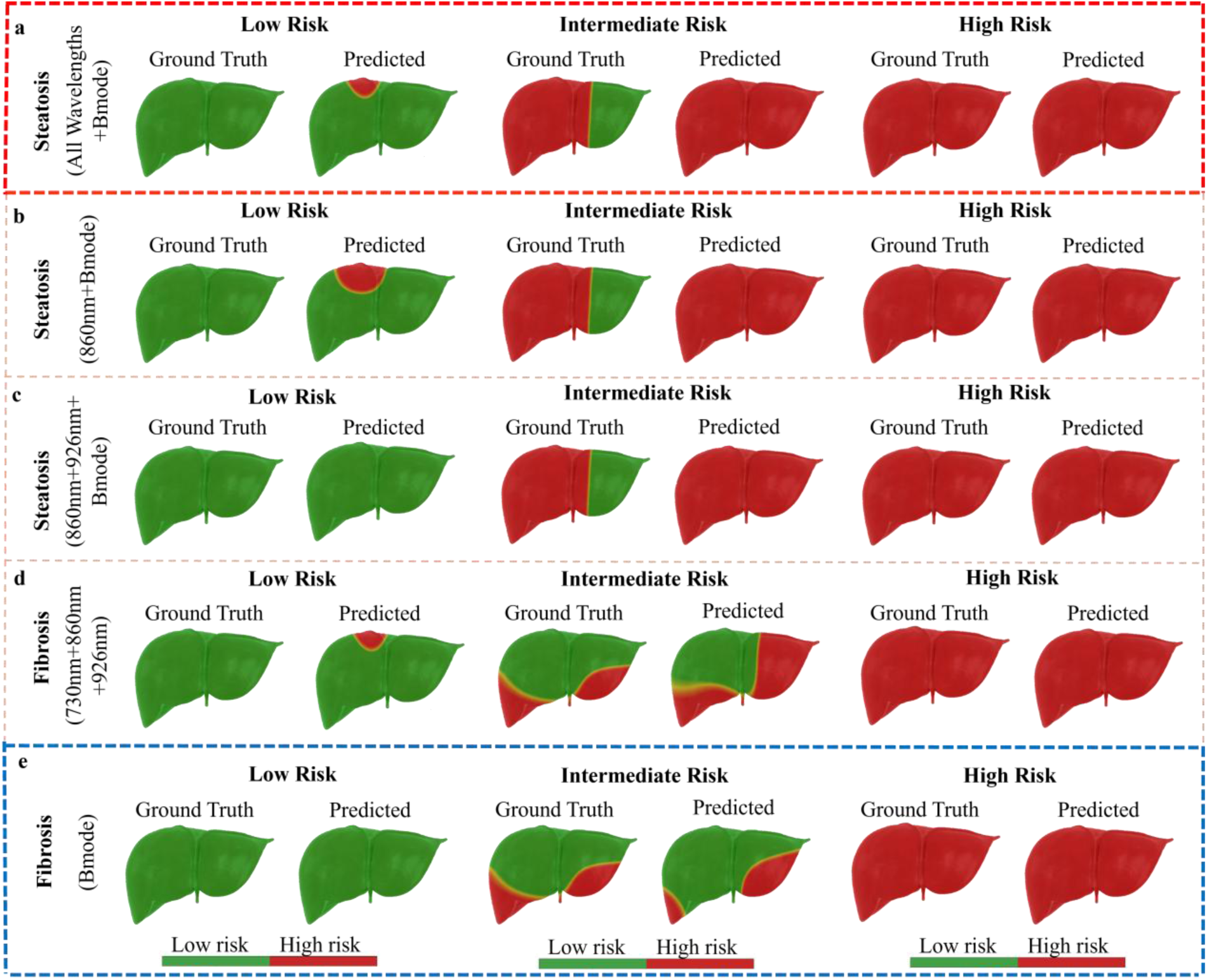
Whole-organ ground truth and predicted risk maps across overall low, intermediate, and high-risk livers for steatosis and fibrosis using different modality combinations. (a) Steatosis (All wavelengths + B-mode). (b) Steatosis (860nm + Bmode). (c) Steatosis (860nm + 926nm + B-mode). (d) Fibrosis (730nm + 860nm +926nm). (e) Fibrosis (Bmode). Green indicates low risk and red indicates high risk.

## Discussion

In this study, we evaluated the feasibility of a deep-learning–assisted multimodal PAT/USI approach for assessing human donor liver quality and transplant suitability using pathology-derived fibrosis and steatosis scores as the reference standard^30, 35, 73, 74^. By combining multispectral PA imaging with B-mode ultrasound, this deep learning model could directly predict liver regional steatosis or fibrosis risk without performing spectral unmixing. In six discarded human donor livers with 60 spatially matched regions of interest, the approach achieved consistent discrimination of steatosis and fibrosis risk levels. Whole-organ prediction maps showed spatial patterns that were generally consistent with histological findings, particularly in intermediate-risk livers where regional heterogeneity was preserved. Together, these findings support a shift from point-based biopsy assessment toward volumetric, image-guided organ evaluation, offering a practical strategy to reduce sampling bias in pre-transplant decision-making^75–80^.

PA and ultrasound contributed differently to steatosis and fibrosis prediction, likely reflecting the different pathological features of these two conditions. For steatosis, multispectral PAT imaging, particularly at 860 nm and 926 nm, contributed substantially to prediction performance. Hepatic steatosis is characterized by lipid accumulation within hepatocytes, which increases optical absorption at lipid-sensitive wavelengths^81–83^. As lipid content increases, optical absorption at lipid-sensitive wavelengths becomes stronger, resulting in increased photoacoustic signal intensity^35^, resulting in improved contrast between low- and high-risk regions and this relationship is further supported by the absorption spectra and phantom validation experiments presented in **Supplementary Fig. 1**. The similar performance observed with reduced wavelength combinations suggested that steatosis-related information is consistently present across multiple PAT channels. In contrast, fibrosis prediction was mainly driven by B-mode ultrasound. Fibrosis is characterized by collagen deposition and architectural remodeling of liver tissue^84^, and these changes predominantly alter acoustic scattering and speckle heterogeneity^85–87^. Therefore, structural ultrasound features are sufficient for fibrosis discrimination^86^, and the addition of PAT information does not substantially improve overall performance^36, 88^.

Conventional multispectral PAT analysis typically relies on spectral unmixing to estimate chromophore concentrations. However, this approach depends on assumptions regarding optical fluence distribution, tissue homogeneity, and predefined absorber spectra, which may not hold in heterogeneous biological tissues such as the liver^36, 41, 88^. Instead of performing explicit spectral unmixing, multispectral PAT volumes were directly used as input to the deep learning model. By learning from the full spatial–spectral information contained in the raw volumetric data, the model bypassed intermediate inversion steps and directly generated predictions of steatosis and fibrosis risk. Such an end-to-end strategy is more clinically practical because it aligns with the goal of pre-transplant evaluation, namely outcome-oriented assessment of organ quality and transplant suitability rather than explicit quantification of individual chromophore concentrations^37, 59, 61^.

Consistent with these methodological considerations, the experimental results further clarified the relative contribution of PAT within the current spectral range. Within the selected wavelength range (680–970 nm), adding PA information did not substantially improve fibrosis prediction compared with B-mode ultrasound alone. This observation may be related to the spectral characteristics of collagen within the current acquisition range^88^. For steatosis, similar performance with fewer wavelengths suggests that lipid-related information is distributed across multiple PA channels. By using the multispectral volumes directly^79, 80, 89^, spatial–spectral information was preserved without relying on explicit inversion procedures. From a practical perspective, reducing the number of wavelengths may shorten acquisition time and simplify system requirements, which is relevant for intraoperative transplant evaluation.

In current transplant practice, frozen-section biopsy samples only a limited portion of the organ and may not fully capture spatial heterogeneity^76–78^, which remains a major challenge in donor liver assessment. An imaging approach capable of providing volumetric risk maps across the whole liver could therefore help identify discordance between local biopsy findings and global tissue condition. Because the proposed method is label-free and relies on rapid ultrasound-based acquisition^73, 74^, it may be integrated into existing evaluation workflows without substantial procedural changes. Rather than replacing histology, such imaging could serve as an adjunct tool to improve confidence in graft acceptance decisions, particularly for marginal organs.

In this study, fibrosis predictions retained regional variation in intermediate-risk livers, suggesting that structural information from ultrasound can be spatially resolved across the organ scale. For steatosis, intermediate livers often contain a large proportion of high-risk regions, which likely contributed to the tendency toward high-risk predictions across broader areas of the organ. All the livers analyzed in this study were declined for clinic transplantation, and many already show extensive fat involvement. As a result, when large areas of an intermediate liver showed high steatosis, the model often predicted high risk across most of the organ. From a clinical perspective, this pattern should be interpreted cautiously. In heterogeneous livers where fat accumulation is widespread, the model may classify most regions as high risk. This tendency highlights the influence of cohort composition on model behavior and underscores the need for validation in more balanced clinical populations^74, 79^.

Several factors should be considered when interpreting these findings in the context of clinical translation. First, the number of donor organs was limited, although multiple spatially matched regions were analyzed within each liver. To mitigate potential overfitting related to within-organ similarity, model evaluation was performed using a leave-one-donor-out strategy, ensuring complete donor-level separation between training and testing. Nevertheless, validation in a larger and more diverse donor cohort remains necessary to confirm generalizability. Second, all livers included in this study were declined for transplantation and many exhibited substantial steatosis. This cohort composition reflects a realistic but imbalanced research scenario and may contribute to the tendency toward higher predicted risk in intermediate livers with widespread fat involvement. Future studies including both accepted and declined grafts will be important to determine how this bias influences clinical decision thresholds. Third, the selected wavelength range (680–970 nm) was optimized for lipid-sensitive contrast but may not fully capture collagen-sensitive spectral features^90–92^. Therefore, the relative contribution of PAT information to fibrosis prediction observed here may depend on the current spectral window and could change with extended near-infrared acquisition.

Future investigations should focus on prospective evaluation across both accepted and declined donor grafts to define clinically meaningful risk thresholds^79^. Beyond transplant evaluation, similar multimodal imaging strategies may also inform noninvasive staging and longitudinal monitoring of metabolic dysfunction–associated steatotic liver disease and advanced fibrosis, where quantitative ultrasound and elastography-based stiffness measurements^93–95^ are widely adopted. In addition, volumetric compositional mapping could support preoperative assessment of background liver quality in patients undergoing major hepatic resection, particularly in the setting of cirrhosis or steatosis-related heterogeneity^96, 97^. Integration with normothermic machine perfusion platforms^98, 99^ may enable longitudinal monitoring of structural and perfusion-related changes during preservation, allowing assessment of dynamic graft recovery or deterioration. Combining PAT-based compositional information with perfusion-sensitive techniques, such as ultrasound localization microscopy^100–102^, may further clarify the relationship between microvascular integrity and tissue pathology^79^. In addition, extending the spectral range toward longer near-infrared wavelengths may improve sensitivity to collagen-related optical signatures and refine fibrosis characterization^91, 92^.

## Conclusion

This study demonstrates the feasibility of a deep-learning–assisted multimodal PAT/USI framework for organ-scale assessment of discarded human donor livers using spatially matched histopathology as the reference standard. By combining multispectral photoacoustic imaging with B-mode ultrasound, the proposed approach directly predicted steatosis and fibrosis risk without relying on spectral unmixing. In this pilot cohort, multispectral PAT contributed most strongly to steatosis discrimination, whereas fibrosis prediction relied primarily on structural ultrasound features. These findings support the potential of volumetric PAT/USI as a rapid and spatially comprehensive adjunct to biopsy for reducing uncertainty in pre-transplant liver evaluation.

## Author contributions

## Data Availability

The data that support the findings of this study are available from the corresponding author upon reasonable request. Due to the use of human donor samples, data sharing may be subject to institutional and ethical regulations.

https://www.dropbox.com/scl/fi/rp689clsva6x3c53p0jow/SupplimentaryMaterials.docx?rlkey=le44dysyalksyort5e0qxenmn&st=v33q1tyl&dl=0

## Acknowledgments

This work was supported by grants from the University of Oklahoma Health Campus (P30CA225520), National Science Foundation (OIA-2132161, 2238648, 2331409), National Institute of Health (R01DK133717), Oklahoma Center for the Advancement of Science and Technology (HR23-071), the medical imaging COBRE (P20 GM135009), the Prevent Cancer Foundation, a grant from the Data Institute for Societal Challenges and the Research Council funded by the Office of the Vice President for Research and Partnerships of the University of Oklahoma Norman Campus, and the Midwest Biomedical Accelerator Consortium (MBArC), an NIH Research Evaluation and Commercialization Hub (REACH). Histology service provided by the Tissue Pathology Shared Resource was supported in part by the National Institute of General Medical Sciences COBRE Grant P20GM103639 and National Cancer Institute Grant P30CA225520 of the National Institutes of Health. Financial support was provided by the OU Libraries’ Open Access Fund.

## Notes

### Competing Interest Statement

The authors have declared no competing interest.

### Funding Statement

This study was supported by grants from the University of Oklahoma Health Sciences Center (P30CA225520), the National Science Foundation (OIA-2132161, 2238648, 2331409), the National Institutes of Health (R01DK133717), the Oklahoma Center for the Advancement of Science and Technology (HR23-071), the Medical Imaging COBRE (P20GM135009), the Prevent Cancer Foundation, the Data Institute for Societal Challenges and the Research Council funded by the Office of the Vice President for Research and Partnerships of the University of Oklahoma Norman Campus, and the Midwest Biomedical Accelerator Consortium (MBArC), an NIH Research Evaluation and Commercialization Hub (REACH).
Histology services were provided by the Tissue Pathology Shared Resource, which is supported in part by the National Institute of General Medical Sciences COBRE Grant (P20GM103639) and the National Cancer Institute Grant (P30CA225520) of the National Institutes of Health.
Open access publication support was provided by the University of Oklahoma Libraries Open Access Fund.

### Author Declarations

The Institutional Review Board of the University of Oklahoma Health Sciences Center gave ethical approval for this work (IRB protocol 12462).

